# CLINICAL, EPIDEMIOLOGICAL AND VACCINATION CHARACTERISTICS IN CHILDREN AND ADOLESCENTS OF SEVERE ACUTE RESPIRATORY SYNDROME DUE TO COVID-19 IN BRAZIL (2020 TO 2024)

**DOI:** 10.1101/2025.09.18.25336058

**Authors:** Vânia Angélica Feitosa Viana, Silvio Alencar Cândido Sobrinho, Francisco de Sousa Júnior, José Quirino da Silva Filho, Karene Ferreira Cavalcante, Débora Bezerra Silva, Leda Maria Simões Mello, Maria Elisabeth Lisboa de Melo, Sônia Maria Santana Macêdo, Shirlene Telmos Silva de Lima, Larissa Maria Façanha Duarte, Fernanda Montenegro Carvalho Araújo, Aldo Ângelo Moreira Lima

**Affiliations:** Post-graduate Program in Medical Sciences, Faculty of Medicine, Federal University of Ceará, Brazil; Central Public Health Laboratory of Ceará, Brazil; NUBIMED - Biomedicine Centre, Faculty of Medicine, Federal University of Ceará, Brazil; University of Campinas, SP, Brazil

**Keywords:** COVID-19, severe acute respiratory syndrome, mortality, case fatality rate

## Abstract

Although less susceptible to severe forms of COVID-19, children and adolescents were also impacted by Severe Acute Respiratory Syndrome (SARS), with hospitalizations and deaths. Brazil recorded one of the highest pediatric mortality rates from COVID-19, exposing structural inequalities and failures in the healthcare system. The objective of this study was to analyze the clinical, epidemiological, and vaccination characteristics of children and adolescents with COVID-19-associated SARS in Brazil (2020–2024). This retrospective observational cohort study analyzed public data from 34,369 SARS cases, revealing disparities in the case fatality rate (CFR): the North (13.02%) and Northeast (11.6%) regions, Indigenous populations (22.54%), rural residents (14.34%), municipalities with a low human development index (9.41%), and the 15–19 age group had higher CFRs. The circulation of the Gamma and Delta variants (2021) was associated with peaks in incidence and mortality; however, during the circulation of Omicron (2022–2024), there was high incidence with reduced mortality. The sustained occupancy of pediatric intensive care units (ICUs) (25–32%) after 2022, and the significant increase in the proportion of children aged 0–4 years in ICU admissions, rising from approximately 52% in 2020–2021 to over 78% in 2024, underscores the demand for targeted strategies for the pediatric population. Comorbidities such as immunosuppression (OR: 4.44) and Down syndrome (OR: 3.13) were independent predictors of death. Vaccination demonstrated a protective effect; however, vaccination coverage was uneven: children under 5 years and the North/Northeast regions had the lowest rates. This study demonstrated that COVID-19-associated SARS in the pediatric population was marked by sociodemographic, regional, clinical, and vaccination disparities. Therefore, there is a clear need for integrated policies that include regionalized strategies for vulnerable populations, strengthening of pediatric ICUs, and equitable expansion of vaccination.

## INTRODUCTION

During the COVID-19 pandemic, although children and adolescents generally presented with milder or asymptomatic clinical presentations compared to adults, severe complications such as Severe Acute Respiratory Syndrome (SARS) were recorded._1_ SARS is characterized as an Influenza-Like Illness (ILI) accompanied by at least one of the following signs of severity: dyspnea/respiratory distress, persistent chest pressure, O_2_ saturation ≤ 94%, and/or cyanosis.^2^ This syndrome emerged as a leading cause of morbidity and mortality in children and adolescents, with a significant number of cases progressing to critical conditions requiring admission to Intensive Care Units (ICUs).^3^

The severity of COVID-19 in children and adolescents is multifactorial, involving pre-existing clinical conditions, age, immunological characteristics, and social determinants. Evidence demonstrates that pediatric hospitalizations for COVID-19 are strongly associated with comorbidities such as chronic cardiopulmonary diseases, immunosuppression, obesity, diabetes, and neurological disorders, which increase the risk of severe outcomes.^4,5^ Furthermore, prematurity, obesity, and elevated inflammatory markers are correlated with a greater need for ICU admission and ventilatory support,^6^ a pattern observed in Brazil, where young infants and children with multiple neurological comorbidities had less favorable prognoses.^7^

It is important to highlight that socioeconomic disparities exacerbate these risks. Limited access to healthcare services, delayed diagnoses, and precarious living conditions act as aggravating factors, exposing vulnerable populations to severe clinical outcomes. This interaction of biological and social factors underscores the need for differentiated approaches for the pediatric population, especially in contexts of structural inequality like Brazil, where profound regional, racial, and socioeconomic inequalities directly influence the epidemiological dynamics and outcomes of COVID-19-associated SARS.^8^ Understanding these variables is essential to inform efficient and equitable public health policies.^9^

Additionally, the virulence of the SARS-CoV-2 variant plays a crucial role in the severity of infection. The high mutagenic capacity of RNA viruses, such as SARS-CoV-2, results in frequent genetic alterations during replication. Although its mutation rate is moderated by the proofreading activity of the NSP14 exonuclease, global dissemination facilitated the emergence of variants with significant impacts on the pandemic. Mutations in the Spike protein, the primary target of the immune response and vaccines, can confer adaptive advantages, such as increased transmissibility, immune evasion, or greater virulence.^10,11^ The trajectory of the pandemic was characterized by epidemic waves related to the emergence of SARS-CoV-2 variants, notably Gamma, Delta, and Omicron, which raised questions about the incidence and mortality of COVID-19 in this age group. Changes in the clinical and epidemiological profile of the disease in the country represent a dynamic challenge for the healthcare system.^12^

The succession of variants in Brazil, with initial predominance of ancestral lineages, followed by the sequential replacement of Gamma, Delta, and Omicron and their sublineages, highlighted the continuous viral evolution and the need for genomic surveillance. Omicron, in particular, underscored the importance of vaccination and the persistent pressure on healthcare systems, even with reduced mortality. This scenario reinforces the relevance of genetic monitoring to guide public policies and immunization strategies, especially in pediatric populations and vulnerable groups.^13^

Vaccination has proven to be a fundamental strategy for reducing severe cases, hospitalizations, and deaths, in addition to enabling a safe return to educational and social activities.^14^ In Brazil, the campaign was implemented in a staggered manner, with ANVISA’s approval of CoronaVac and Pfizer-BioNTech (BNT162b2) for different age groups starting in 2021. In February 2021, vaccination began for those aged 16 and over, expanding in June of the same year to the 12-16 age group. In January 2022, vaccination started for ages 5-11, and in September, for ages 6 months to 5 years.^15,1,17^

Vaccination coverage in the pediatric population in Brazil remains insufficient. As of April 2025, 65.43% of the 53.6 million individuals aged 0-19 had received two doses, but only 29.08% had taken the third dose.^18^ Vaccine hesitancy, misinformation, and underestimation of the severity of COVID-19 in childhood contributed to this low adherence, jeopardizing lasting and collective protection.^19^ The incorporation of the vaccine into the National Immunization Calendar in 2024 for children aged 6 months to 5 years represents an advance in protecting this group and reinforces the commitment to public health and equity.^20^

By encompassing a five-year period (2020-2024), this research provides a novel and necessary overview that contributes to the understanding of the dynamics of COVID-19-associated SARS in this population. In light of the above, it is pertinent to elucidate the determinants of morbidity, mortality, and lethality from COVID-19 in the 0-19 age group through an analysis integrating clinical, epidemiological, and vaccination data from all regions of Brazil. Studying the factors associated with death from SARS allows for the direction of prevention strategies, early diagnosis, and clinical management tailored to this population.

## METHODS

### Ethical aspects

The data used in this study are publicly available, obtained from secondary and anonymized sources, in compliance with the Brazilian Access to Information Act (Law No. 12,527/2011). In accordance with CNS Resolution No. 510/2016, research with these characteristics is exempt from evaluation by a Research Ethics Committee (REC) and from the requirement of obtaining Informed Consent Forms (ICF).

### Study design

This is an observational, retrospective cohort study conducted through the analysis of aggregated secondary data on reported cases of Severe Acute Respiratory Syndrome (SARS) due to COVID-19 in children and adolescents (0 to 19 years) in Brazil. The analyzed period spans from January 1, 2020, to December 31, 2024, corresponding to the first five years of SARS-CoV-2 virus circulation.

### Data source

Data on SARS cases due to COVID-19 in children and adolescents were obtained from the Influenza Epidemiological Surveillance Information System (SIVEP-Gripe), available on the OpenDataSUS platform, which contains sociodemographic, clinical, epidemiological, and vaccination information. Detailed vaccination data (doses, timing, manufacturer) were supplemented with records from the National Vaccination Campaign database (OpenDataSUS).

The characterization of SARS-CoV-2 lineages was based on the Fiocruz Genomic Network repository^21^. The Municipal Human Development Index (MHDI), used as a proxy for socioeconomic context, was obtained from the United Nations Development Programme^22^, and stratified by interquartile intervals.

Population estimates by age group and municipality, used for calculating adjusted rates, were retrieved from the IBGA Automatic Recovery System (SIDRA)^23^.

### Study selection criteria

All cases of children and adolescents with SARS associated with COVID-19 recorded in the SARS Database – including COVID-19 data (OpenDataSUS) – that met the criteria outlined in Figure 1 were included.

**Figure 1.**
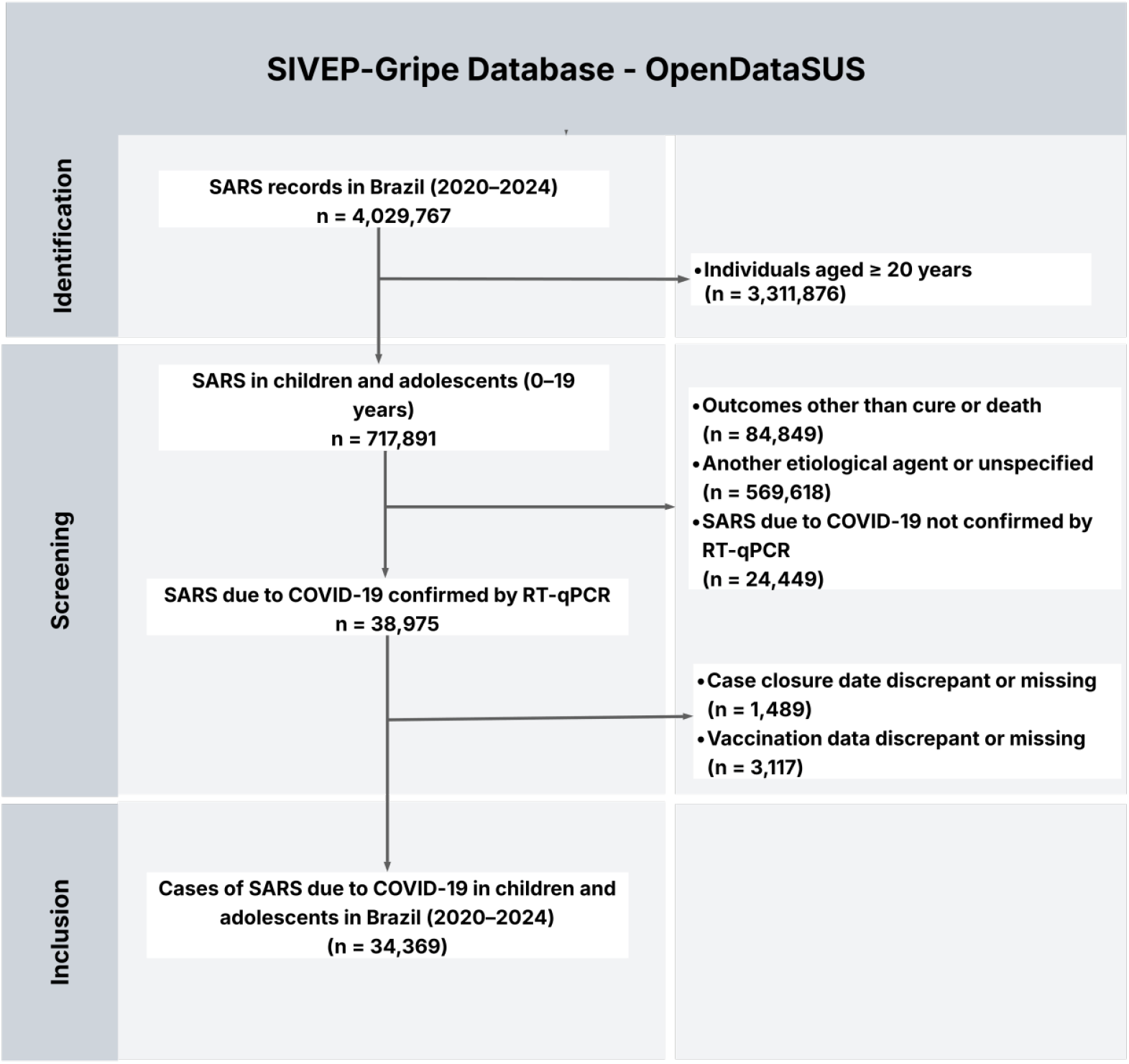
Selection process of cases of Severe Acute Respiratory Syndrome (SARS) due to COVID-19 in children and adolescents in Brazil, 2020 to 2024. Source: Prepared by the author(s).

### Data processing and statistical analysis

Initial data processing was performed using Bash, AWK, and R (R Core Team, 2025). Graphical visualizations were generated using packages from R and Python.

Comparisons between proportions employed the Chi-square (χ^2^) test and Z-test for Case Fatality Rates (CFR), utilizing the rstatix and rcompanion packages in the R programming language.^24,25^

Incidence and mortality rates were age-adjusted based on the 2022 Census (using the sidrar package).^23^ The Case Fatality Rate was calculated as the ratio between the number of deaths from SARS associated with COVID-19 and the total number of SARS cases, multiplied by 100.^26^

Vaccination status was categorized as follows: unvaccinated: individuals who did not receive any dose of the COVID-19 vaccine; 1st/2nd dose: those who received only the first dose or both doses of the primary vaccination schedule and 1st/2nd booster: individuals who received at least one booster dose.

Based on this categorization, two complementary measures were estimated: the proportion of vaccinated individuals and booster dose adherence. The proportion of vaccinated individuals was defined as the percentage of children and adolescents with severe COVID-19 who were vaccinated relative to the total study population with SARS. Booster adherence, in turn, was defined as the proportion of individuals who received at least one additional dose among those previously vaccinated. The vaccinated group was considered the union of individuals in the ‘1st/2nd dose’ and ‘1st/2nd booster’ categories.

The association between variables and the outcome of death was estimated using Odds Ratios (OR) and their 95% Confidence Intervals (95% CI), employing binomial logistic regression models. An OR > 1 indicates a higher chance of death, OR < 1 suggests a protective effect, and OR = 1 indicates no association. The analyses were restricted to cases with complete information (n=21,817).

Univariate Generalized Linear Models (GLMs) were fitted for sociodemographic variables; signs and symptoms; comorbidities; ICU admission, use of ventilatory support, and vaccination. The final multivariate model was constructed using stepwise regression (MASS package), including variables with p < 0.05 and OR > 1.0 in the univariate analysis, in addition to sociodemographic and vaccination variables of interest.^27^

## RESULT

### Sociodemographic characteristics and Case Fatality Rate (CFR) analysis

The analysis revealed an overall case fatality rate (CFR) of 5.9% during the study period, with a heterogeneous distribution among population subgroups (p < 0.05). The highest rates were associated with the North (13.02%) and Northeast (11.6%) regions; the 15 to 19-year age group (10.24%); the female sex (6.24%); municipalities with a low Human Development Index (9.41%); Indigenous race/color (22.54%); and rural residence (14.34%). These results indicate higher fatality rates among historically vulnerable populations and in regions with lower socioeconomic indicators (Table 1).

**Table 1.** Sociodemographic characteristics and Case Fatality Rate (CFR) analysis of Severe Acute Respiratory Syndrome (SARS) COVID-19 cases in children and adolescents, Brazil, 2020 to 2024.

| Category | Factor Level | Cure n(%) | Death n(%) | CFR (%) | $\chi^2$ p-value |
| --- | --- | --- | --- | --- | --- |
| **REGION (%)** | **Central-West** | 3710 (11.5) | 178 ( 8.8) | 4.58b | **<0.001** |
|  | **Northeast** | 5169 (16.0) | 678 (33.4) | 11.6a |  |
|  | **North** | 1516 ( 4.7) | 227 (11.2) | 13.02a |  |
|  | **Southeast** | 15785 (48.8) | 691 (34.0) | 4.19b |  |
|  | **South** | 6158 (19.0) | 257 (12.7) | 4.01b |  |
| **AGE_YEAR (%)** | **0 - 4** | 20541 (63.5) | 1061 (52.2) | 4.91c | **<0.001** |
|  | **5 - 9** | 4092 (12.7) | 212 (10.4) | 4.93c |  |
|  | **10 -14** | 3048 ( 9.4) | 227 (11.2) | 6.93b |  |
|  | **15 - 19** | 4657 (14.4) | 531 (26.1) | 10.24a |  |
| **HDI_2010 (%)** | **0.805 - 0.862** | 12245 (38.7) | 399 (20.0) | 3.16d | **<0.001** |
|  | **0.784 - 0.804** | 5283 (16.7) | 292 (14.6) | 5.24c |  |
|  | **0.752 - 0.783** | 7754 (24.5) | 643 (32.2) | 7.66b |  |
|  | **0.418 - 0.751** | 6353 (20.1) | 660 (33.1) | 9.41a |  |
| **SEX (%)** | **Female** | 14863 (46.0) | 990 (48.8) | 6.24a | **0.015** |
|  | **Male** | 17459 (54.0) | 1039 (51.2) | 5.62b |  |
| **RACE (%)** | **White** | 12903 (50.3) | 644 (37.0) | 4.75c | **<0.001** |
|  | **Mixed/Parda** | 11533 (45.0) | 989 (56.8) | 7.9b |  |
|  | **Black** | 895 ( 3.5) | 59 ( 3.4) | 6.18bc |  |
|  | **Asian** | 186 ( 0.7) | 11 ( 0.6) | 5.58bc |  |
|  | **Indigenous** | 134 ( 0.5) | 39 ( 2.2) | 22.54a |  |
| **AREA (%)** | **Urban** | 27449 (94.5) | 1609 (88.4) | 5.54b | **<0.001** |
|  | **Peri-urban** | 446 ( 1.5) | 17 ( 0.9) | 3.67b |  |
|  | **Rural** | 1159 ( 4.0) | 194 (10.7) | 14.34a |  |
| **TOTAL** |  | **32338** | **2031** | **5.9** |  |

Source: Prepared by the authors using data from SIVEP-Gripe/OpenDataSUS, 2020-2024.
Note: Comparisons between CFRs were performed using the Chi-square and Z-tests. Letters (a, b, c, and d) indicate differences between groups. Proportions sharing the same letter do not differ significantly, whereas distinct letters represent statistically different groups ( $p < 0.05$ ). Values are sorted in descending order (from the highest to the lowest proportion). The total number of cases per variable may differ from the total sample size ( $n=34,369$ ) due to missing data (information not recorded in the notification form).

### Temporal distribution of cases, deaths, and case fatality rate of COVID-19-associated SARS in children and adolescents

Of the analyzed period, the year with the highest number of COVID-19-associated SARS cases in children and adolescents was 2022 (n=10,096), followed by 2021 (n=9,335) and 2020 (n=7,286). From 2023 onwards, a progressive reduction was observed, with 4,204 SARS cases in 2023 and 3,448 in 2024, indicating a declining trend in COVID-19-associated SARS occurrences.

Regarding deaths (n=2,031), the highest number was recorded in 2021 (n=733), followed by 2020 (n=673). From 2022 onwards, a significant drop in deaths was observed. The CFR also showed a reduction over the five years, starting from 9.24% in 2020 to 2.99% in 2024.

The temporal evolution of the pandemic revealed that the highest peak in lethality occurred at the beginning of the pandemic, with 12.3% in the second quarter of 2020, followed by 11.1% in the first quarter of 2020 and 9.5% in the first quarter of 2021 (2021-Q1). After 2022, a sustained downward trend was observed, with the CFR stabilizing at levels between 2% and 4%, even with the absolute peak of cases (>5,000) recorded in the first quarter of 2022. The number of deaths declined progressively, remaining below 500 per quarter from the second quarter of 2021 onwards (Figure 2).

**Figure 2.**
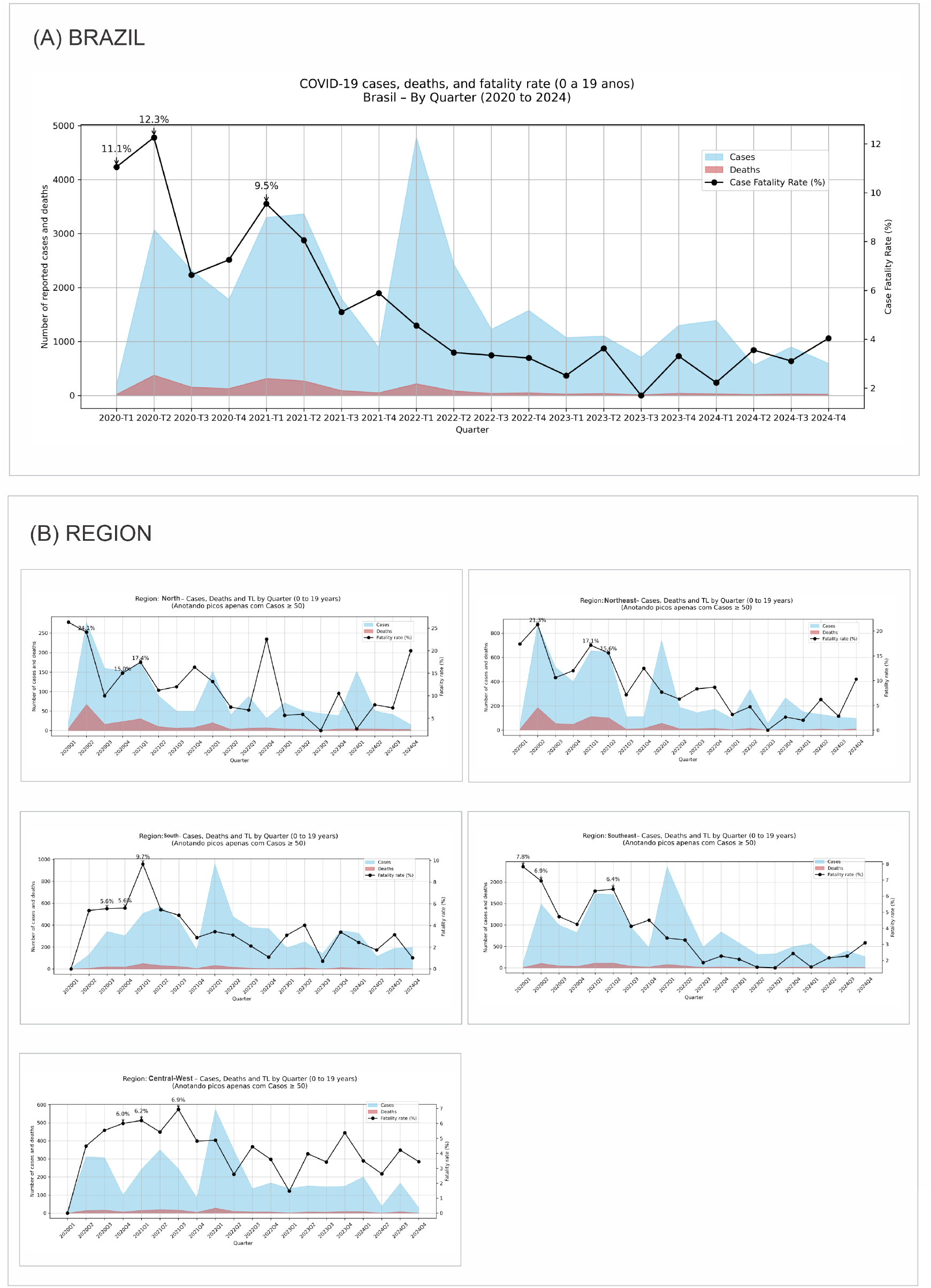
Cases and deaths from Severe Acute Respiratory Syndrome (SARS) due to COVID-19 and the Case Fatality Rate (CFR) among children and adolescents, by year of notification and region, Brazil, 2020 to 2024. Source: Prepared by the authors using data from SIVEP-Gripe/OpenDataSUS, 2020-2024. Note: The CFR was calculated as (number of deaths / number of cases) × 100. The North and Northeast regions had the highest CFR throughout the period. The data refer to the full five-year period, from 2020 to 2024. T1, T2, T3, and T4 refer to the 1st, 2nd, 3rd, and 4th quarters, respectively.Factors associated with death from COVID-19 in children and adolescents with SARS.

Regional disparities were striking and persistent. The North region presented the highest and most unstable rates, with extreme peaks at the beginning of the pandemic (>26% in Q1 2020) and new outbreaks of high lethality (>20%) still in the fourth quarter of 2022 and the fourth quarter of 2024. A similar, though less intense, pattern was observed in the Northeast, which recorded a CFR of 21.3% in Q2 2020 and 10% in Q4 2024 (Figure 2).

In contrast, the Southeast and South regions exhibited more controlled patterns, with their highest rates at the beginning of the pandemic (a maximum of 9.7% in the South in Q1 2021) and rapid stabilization around 2% to 4% from 2022 onwards. The Central-West region maintained intermediate and relatively constant rates, with a peak of 6.9% in Q3 2021 (Figure 2).

Regarding age distribution, the 15 to 19-year-old age group consistently presented the highest lethality rate every year (11.2% in 2020; 11.3% in 2021; 7.5% in 2024). Although the 0 to 4-year-old age group accounted for the highest absolute number of cases, its CFR was significantly lower and showed the most significant reduction, falling from 7.5% in 2020 to 2.7% in 2024.

In summary, the results demonstrate a clear transition from an acute phase with extremely high lethality to an endemic scenario with controlled rates, yet with persistent regional and age-related inequities. The North and Northeast regions and adolescents (15-19 years) remain the most vulnerable groups.

### Factors associated with death from COVID-19 in children and adolescents with SARS

Of the 2,031 deaths analyzed, 62.1% (n=1,261) occurred in individuals with at least one comorbidity. The most frequent were neurological disease (24.5%), cardiopathy (19.7%), and immunosuppression (14.1%). A distinct age profile was observed: cardiopathies, Down syndrome, and pneumopathies predominated in children <10 years, while obesity (90.7%) and diabetes (64.5%) were more frequent among adolescents (10-19 years). The clinical presentation was predominantly respiratory, with dyspnea (71.2%), respiratory distress (68.1%), and O_2_ saturation <95% (64.6%) being the most common signs.

Univariate analysis identified strong associations between death and sociodemographic factors, such as residence in the North (OR: 2.86) and Northeast (OR: 2.61) regions, rural area (OR: 2.78), low municipal HDI (OR: 2.89), Indigenous race (OR: 4.91), and the 15-19 year age group (OR: 2.25). Among comorbidities, chronic liver disease (OR: 5.34) and immunosuppression (OR: 4.69) presented the highest odds for the outcome. Vaccination showed a significant protective effect (OR<1; p<0.005).

In the multivariate analysis, several factors remained as independent predictors of death. Higher odds of death were associated with the following sociodemographic factors: residence in the Northeast region (aOR: 2.28), self-reported Indigenous race (aOR: 2.80), age 15-19 years (aOR: 2.55), rural residence (aOR: 1.62), and low municipal human development index (aOR: 2.06). Among comorbidities, the strongest predictors were immunosuppression (aOR: 4.44), Down syndrome (aOR: 3.13), and chronic liver disease (aOR: 3.16), followed by cardiopathy (aOR: 1.96) and neurological disease (aOR: 1.94). Clinical signs of severity and care variables also showed significant associations: O_2_ saturation <95% (aOR: 1.37), ICU admission (aOR: 1.76), and, most notably, the need for invasive ventilatory support (aOR: 22.34). Conversely, vaccination provided robust protection, reducing the odds of death by more than 50%; this effect was more pronounced with at least one booster dose (for 1st/2nd dose: aOR: 0.46; with booster: aOR: 0.29) (Figure 3).

**Figure 3.**
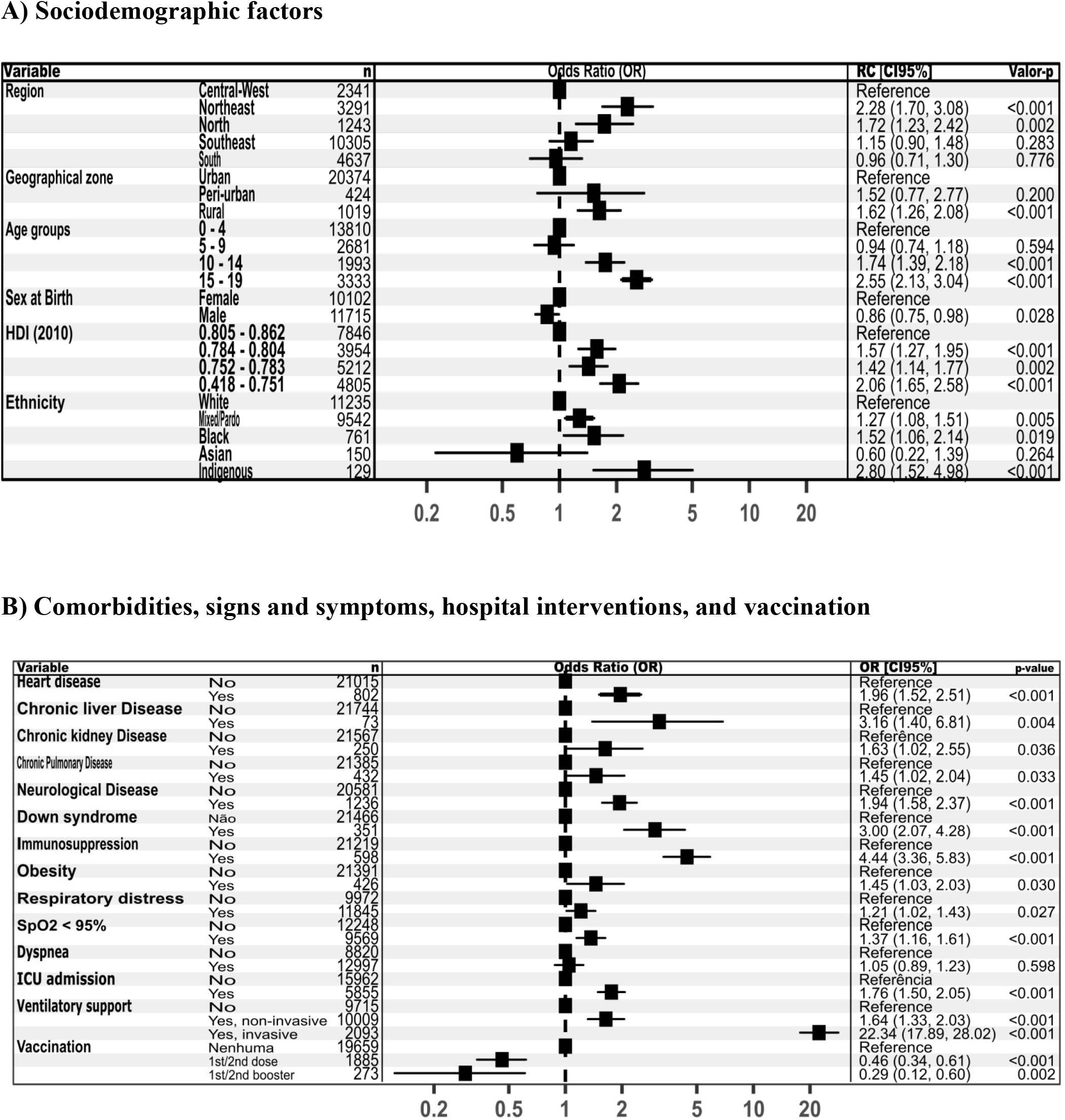
Multivariate analysis of factors associated with death from COVID-19 in children and adolescents with Severe Acute Respiratory Syndrome (SARS), Brazil, 2020 to 2024. Source: Prepared by the authors using data from SIVEP-Gripe/OpenDataSUS, 2020-2024. Chart derived from a Generalized Linear Model (GLM) with binomial distribution, performed only with complete data (n=21,817). OR > 1 indicates a higher chance of death; OR < 1 suggests a protective factor and OR=1 indicates no association.

In summary, the outcome of death was associated with a complex set of sociodemographic inequities, specific comorbidities, clinical signs of severity, and the need for ventilatory support, while vaccination proved to be a fundamental protective measure.

Furthermore, the analysis of ICU admissions revealed an age-related shift in case severity over time. Of the total 9,139 ICU admissions recorded (26.6% of all cases), the year 2022 accounted for the highest absolute number (n=2,557). A significant and increasing trend was observed in the proportion of ICU admissions among the 0 to 4-year-old age group. This group, which represented approximately 52% of ICU admissions in 2020 and 2021, came to comprise the majority of cases starting in 2022, reaching 78.6% in 2024 (763 out of 971 admissions) (Table 2).

**Table 2.** Distribution of cases of Severe Acute Respiratory Syndrome (SARS) due to COVID-19 and Intensive Care Unit (ICU) admissions among children and adolescents, according to age group and year of notification, Brazil, 2020-2024.

| **Year of Notification** | **Age Group (years)** | **Total SARS n(%)** | **ICU/Age Group n(%)** | **ICU/Year %** |
| --- | --- | --- | --- | --- |
| **2020** | **0 - 4** | 3,598 (49.4) | 938 (26.1) | 52.6 |
|  | **5 - 9** | 1,052 (14.4) | 237 (22.5) | 13.3 |
|  | **10 - 14** | 942 (12.9) | 245 (26.0) | 13.7 |
|  | **15 - 19** | 1,694 (23.3) | 363 (21.4) | 20.4 |
|  | **Total** | **7,286 (100)** | **1,783** | **100** |
| **2021** | **0 - 4** | 4,739 (50.8) | 1,342 (28.3) | 51.7 |
|  | **5 - 9** | 1,101 (11.8) | 275 (25.0) | 10.6 |
|  | **10 - 14** | 1,150 (12.3) | 339 (29.5) | 13.1 |
|  | **15 - 19** | 2,345 (25.1) | 639 (27.2) | 24.6 |
|  | **Total** | **9,335 (100)** | **2,595** | **100** |
| **2022** | **0 - 4** | 7,157 (70.9) | 1,857 (25.9) | 72.6 |
|  | **5 - 9** | 1,273 (12.6) | 295 (23.2) | 11.5 |
|  | **10 - 14** | 803 (8.0) | 188 (23.4) | 7.4 |
|  | **15 - 19** | 863 (8.5) | 217 (25.1) | 8.5 |
|  | **Total** | **10,096 (100)** | **2,557** | **100** |
| **2023** | **0 - 4** | 3,358 (79.9) | 961 (28.6) | 77.9 |
|  | **5 - 9** | 454 (10.8) | 146 (32.2) | 11.8 |
|  | **10 - 14** | 226 (5.4) | 72 (31.9) | 5.8 |
|  | **15 - 19** | 166 (3.9) | 54 (32.5) | 4.4 |
|  | **Total** | **4,204 (100)** | **1,233** | **100** |
| **2024** | **0 - 4** | 2,750 (79.7) | 763 (27.7) | 78.6 |
|  | **5 - 9** | 424 (12.3) | 129 (30.4) | 13.3 |
|  | **10 - 14** | 154 (4.5) | 46 (29.9) | 4.7 |
|  | **15 - 19** | 120 (3.5) | 33 (27.5) | 3.4 |
|  | **Total** | **3,448 (100)** | **971** | **100** |

Source: Prepared by the authors using data from SIVEP-Gripe/OpenDataSUS, 2020-2024.
Note: ICU/Age group - Percentage of ICU admissions by age group. ICU/Year - Percentage of ICU admissions by year of notification.

In contrast, when analyzing the ICU admission rate per case within each age group, the pattern was distinct. While children aged 0 to 4 years maintained a stable rate (25-29%), the 5-9, 10-14, and 15-19-year-old age groups exhibited a marked increase in severity starting in 2023, with ICU admission rates exceeding 30% in each group (Table 2). This finding indicates that although children under 5 became numerically predominant in the ICUs, cases among older children and adolescents became proportionally more severe in the final year of the study.

### Proportion of children and adolescents with SARS vaccinated against COVID-19, by age group and region, Brazil

The analysis of vaccination status revealed low vaccine uptake among the 34,369 cases of COVID-19-associated SARS. The vast majority of cases (90%) had not received any dose of the vaccine, a percentage that was even higher among the 2,031 deaths (94.1%). Only a small minority had received one or two doses (8.7% of cases; 5.4% of deaths) or at least one booster dose (1.2% of cases; 0.5% of deaths) (Table 3).

**Table 3.** Proportion of children and adolescents with Severe Acute Respiratory Syndrome (SARS) vaccinated against COVID-19 and booster dose adherence, by region and age group, Brazil, 2020-2024.

| Category | Factor Level | Total SARS | Unvaccinated n(%) | 1st/2nd dose n(%) | 1st/2nd booster n(%) | Proportion of vaccinated* (%) | Booster adherence ** (%) |
| --- | --- | --- | --- | --- | --- | --- | --- |
| Region | Central-West | 3,888 | 3,532 (90.8) | 315 (8.1) | 41 (1.0) | 9.20 | 11.50 |
|  | Northeast | 5,847 | 5,334 (91.2) | 450 (7.6) | 63 (1.0) | 9.60 | 12.30 |
|  | North | 1,743 | 1,630 (93.5) | 103 (5.9) | 10 (0.5) | 6.50 | 8.80 |
|  | Southeast | 16,476 | 14,687 (89.1) | 1,554 (9.4) | 235 (1.4) | 10.90 | 13.10 |
|  | South | 6,415 | 5,759 (89.7) | 580 (9.0) | 76 (1.1) | 10.20 | 11.60 |
| Age Group | 0-4 years | 21,602 | 20,733 (95.9) | 752 (3.4) | 117 (0.5) | 4.00 | 13.50 |
|  | 5-9 years | 4,304 | 3,461 (80.4) | 805 (18.7) | 38 (0.8) | 19.60 | 4.50 |
|  | 10-14 years | 3,275 | 2,596 (79.2) | 603 (18.4) | 76 (2.3) | 20.70 | 11.20 |
|  | 15-19 years | 5,188 | 4,152 (80.0) | 842 (16.2) | 194 (3.7) | 20.00 | 18.70 |

Source: Prepared by the authors using data from SIVEP-Gripe/OpenDataSUS, 2020-2024.
Note: \*Vaccination proportion = (1<sup>st</sup>/2<sup>nd</sup> dose + 1<sup>st</sup>/2<sup>nd</sup> booster) / Total SARS cases × 100.
\*\*Booster adherence = (1<sup>st</sup>/2<sup>nd</sup> booster) / (1<sup>st</sup>/2<sup>nd</sup> dose + 1<sup>st</sup>/2<sup>nd</sup> booster) × 100.

Disparities in vaccination status were evident when stratified by age group and region. Children aged 0 to 4 years, who constituted the group with the highest absolute number of cases (n=21,602), also had the highest proportion of unvaccinated individuals (95.9%). In contrast, adolescents (10-19 years) recorded the highest proportions of vaccination, notably the 15-19-year-old group, which had the highest uptake of the booster dose (18.7%). Geographically, the North (93.5%) and Northeast (91.2%) regions had the highest proportions of unvaccinated individuals, while the Southeast region stood out with the highest proportions of individuals who had received at least one dose (10.9%) and at least one booster (13.1%) (Table 3).

These results demonstrate a strong association between non-vaccination and the occurrence of severe cases and deaths, with significant inequities in vaccine protection related to age and geographic region.

### Impact of SARS-CoV-2 variants and COVID-19 vaccination on the incidence and mortality from Severe Acute Respiratory Syndrome (SARS) in children and adolescents in Brazil

The temporal analysis revealed a clear transition in the epidemiological profile of COVID-19-associated SARS in children and adolescents, closely associated with the emergence of SARS-CoV-2 variants and the implementation of vaccination. The initial peaks in incidence and mortality (2020–2021) coincided with the circulation of the Gamma and Delta variants, marking a period of higher severity. Starting in 2022, the spread of the Omicron variant and its sublineages led to the most significant peak in incidence throughout the historical series, yet with a sharp reduction in mortality, a trend that continued with low and stable levels in 2023 and 2024 (Figure 4).

**Figure 4.**
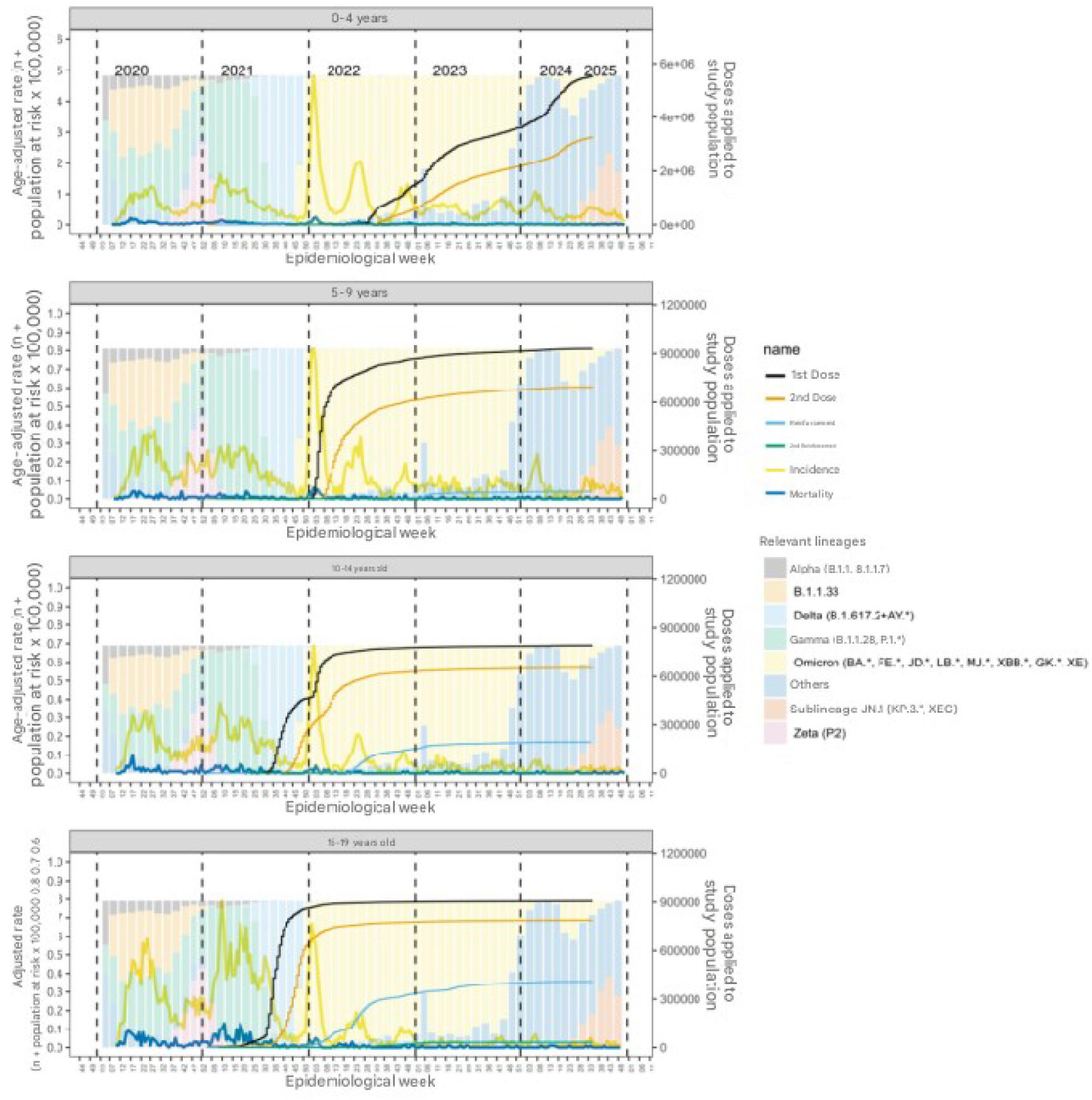
Incidence and mortality rates in children and adolescents by epidemiological week and age group, circulating SARS-CoV-2 variants, and COVID-19 vaccination, Brazil, 2020 to 2024. Source: Prepared by the authors using data from SIVEP-Gripe, the National COVID-19 Vaccination Campaign, and the Fiocruz Genomic Network, 2020-2024. Note: Age-adjusted incidence and mortality rates of SARS cases (left y-axis) by epidemiological week (x-axis), relative frequency of circulating variants (background bars scaled from 0 to 100% of the maximum relative frequency), and vaccination rollout across different age groups, Brazil, from January 1, 2020, to December 31, 2024 (5 years).

The impact differed substantially between age groups. Children aged 0 to 4 years, the last group to be vaccinated (2022), exhibited the highest case volumes and the most significant mortality rates in the pre-vaccination period. Following the introduction of the vaccine, a clear decrease in these indicators was observed. A similar pattern, though with lower magnitudes of incidence and mortality, was seen in the 5 to 9-year-old group (Figure 4).

Among adolescents aged 10 to 14 years, vaccination initiated in the second half of 2021 was followed by a sustained decline in mortality, even during the Omicron case peak. The 15 to 19-year-old group stood out for having the highest mortality rate early in the pandemic, during the Gamma and Delta waves. The rapid vaccination of this group (2021–2022) was crucial in decoupling high Omicron transmissibility from fatal outcomes, resulting in incidence peaks without a return to previous mortality levels (Figure 4).

In summary, the data demonstrate that vaccination was a determining factor in decoupling incidence from mortality, significantly mitigating the impact of more transmissible variants and reducing the disease burden most markedly in age groups with higher vaccine uptake.

## DISCUSSION

These results outline a multifactorial landscape of severe COVID-19 outcomes in the Brazilian pediatric population, in which social and public health determinants played a crucial role. The disproportionate burden of the disease among Indigenous children and adolescents, those residing in the North and Northeast regions, in rural areas, and in municipalities with a low HDI reflects historical inequities in access to healthcare services and structural socioeconomic vulnerabilities.^19,28^ This pattern of disparity, consistent throughout the entire study period, was confirmed as an independent predictor of death in the multivariate analysis, demonstrating that socioeconomic conditions and regional location are as determinant for the outcome as clinical factors.

The temporal analysis revealed the evolution of the pandemic in distinct phases. The peak incidence in 2022, coinciding with the circulation of the Omicron variant, contrasted with the higher mortality recorded in 2020–2021, a period marked by the Gamma and Delta variants and the absence of specific immunization.^29,30^ The sustained decline in both incidence and mortality from 2022 onward suggests an epidemiological transition influenced by the combination of two main factors: the expansion of vaccination coverage and the possible intrinsic reduction in the virulence of circulating viral lineages^.31,32^

The study corroborates the established literature on the profile of comorbidities that increase the risk of fatal outcomes, with immunosuppression, Down syndrome, and chronic liver diseases emerging as the highest-risk conditions.^33,34^ Severe clinical presentation upon admission, characterized by low oxygen saturation and respiratory distress, and the need for ICU admission and invasive ventilatory support remained strong independent predictors of death, underscoring the severity of these patients.^29^

One of the most significant findings was the graded and robust protective effect of vaccination, with at least one booster dose being associated with the most substantial reduction in the odds of death. This finding is particularly relevant given the low vaccination coverage observed in younger children and in the most vulnerable regions of the country, pointing to the urgency of targeted campaigns for these groups.^18^

In summary, the findings of this study reinforce that the trajectory of severe COVID-19 in children and adolescents in Brazil was profoundly modulated by social determinants. The attenuation of mortality observed in the most recent period is closely linked to public health interventions, notably vaccination. Therefore, future policies must integrate the continuous strengthening of pediatric immunization, especially among vulnerable populations, with the enhancement of the healthcare network to ensure equitable and timely access to care, preparing the system for future health emergencies.

## CONCLUSIONS

This national analysis of COVID-19-associated SARS in children and adolescents from 2020 to 2024 reveals critical insights into the evolving epidemiology of the disease and the impact of public health interventions.

A heterogeneous case fatality rate was observed, significantly higher in the North and Northeast regions, among Indigenous populations, rural residents, municipalities with a low Human Development Index (HDI), and adolescents aged 15 to 19 years. Multivariate analysis confirmed that these sociodemographic and geographic disparities are independent risk factors for death, alongside pre-existing clinical conditions (immunosuppression, liver diseases, Down syndrome, cardiopathies, and obesity) and severe clinical presentation upon admission (SpO_2_ < 95%, respiratory distress, ICU admission, and need for ventilatory support).

The temporal evolution of incidence and mortality was marked by two distinct phases. The peak incidence occurred in 2022, while mortality was highest in 2020, followed by a significant and sustained decline starting in 2022. This pattern coincided with the emergence of the Omicron variant, associated with higher transmissibility but lower severity, and crucially, with the rollout of the COVID-19 vaccination campaign. Vaccination, particularly with at least one booster dose, was identified as a protective factor, underscoring its importance even in age groups with lower inherent risk, despite lower vaccination coverage among children aged 0 to 4 years and in the North and Northeast regions.

Therefore, these findings highlight the impact of vaccination in reducing pediatric mortality and the persistent inequality in health outcomes determined by social factors. This reinforces the urgent need for integrated public health policies that prioritize: equitable vaccination strategies targeting vulnerable populations and regions with lower coverage; strengthening pediatric ICU capacity; and regionalized approaches to address the specific socioeconomic vulnerabilities that exacerbate the spread and severity of infectious diseases. These measures are essential to mitigate pediatric morbidity and mortality in future health emergencies.

## Data Availability

All data produced in the present work are contained in the manuscript.

## ACKNOWLEDGEMENTS

The authors thank Brazilian public institutions for providing and maintaining the open data platforms that made this study possible.

